# Wastewater and surface monitoring to detect COVID-19 in elementary school settings: The Safer at School Early Alert project

**DOI:** 10.1101/2021.10.19.21265226

**Authors:** Rebecca Fielding-Miller, Smruthi Karthikeyan, Tommi Gaines, Richard S. Garfein, Rodolfo A. Salido, Victor J. Cantu, Laura Kohn, Natasha K Martin, Adriane Wynn, Carrissa Wijaya, Marlene Flores, Vinton Omaleki, Araz Majnoonian, Patricia Gonzalez-Zuniga, Megan Nguyen, Anh V Vo, Tina Le, Dawn Duong, Ashkan Hassani, Samantha Tweeten, Kristen Jepsen, Benjamin Henson, Abbas Hakim, Amanda Birmingham, Peter De Hoff, Adam M. Mark, Chanond A Nasamran, Sara Brin Rosenthal, Niema Moshiri, Kathleen M. Fisch, Greg Humphrey, Sawyer Farmer, Helena M. Tubb, Tommy Valles, Justin Morris, Jaeyoung Kang, Behnam Khaleghi, Colin Young, Ameen D Akel, Sean Eilert, Justin Eno, Ken Curewitz, Louise C Laurent, Tajana Rosing, Rob Knight

## Abstract

**Background:** Schools are high-risk settings for SARS-CoV-2 transmission, but necessary for children’s educational and social-emotional wellbeing. Previous research suggests that wastewater monitoring can detect SARS-CoV-2 infections in controlled residential settings with high levels of accuracy. However, its effective accuracy, cost, and feasibility in non-residential community settings is unknown.

**Methods:** The objective of this study was to determine the effectiveness and accuracy of community-based passive wastewater and surface (environmental) surveillance to detect SARS-CoV-2 infection in neighborhood schools compared to weekly diagnostic (PCR) testing. We implemented an environmental surveillance system in nine elementary schools with 1700 regularly present staff and students in southern California. The system was validated from November 2020 – March 2021.

**Findings:** In 447 data collection days across the nine sites 89 individuals tested positive for COVID-19, and SARS-CoV-2 was detected in 374 surface samples and 133 wastewater samples. Ninety-three percent of identified cases were associated with an environmental sample (95% CI: 88% - 98%); 67% were associated with a positive wastewater sample (95% CI: 57% - 77%), and 40% were associated with a positive surface sample (95% CI: 29% - 52%). The techniques we utilized allowed for near-complete genomic sequencing of wastewater and surface samples.

**Interpretation:** Passive environmental surveillance can detect the presence of COVID-19 cases in non-residential community school settings with a high degree of accuracy.

**Funding:** County of San Diego, Health and Human Services Agency, National Institutes of Health, National Science Foundation, Centers for Disease Control

## Background

Safely operating schools during the COVID-19 pandemic is a public health challenge. In schools, unvaccinated individuals spend extended amounts of time in close proximity, typically indoors. They are therefore potentially high-risk spaces for respiratory virus transmission.

Minimizing learning loss due to illness is essential for children’s social, physical, and emotional wellbeing^1^. Additionally, caretaking responsibilities for children experiencing either acute illness or the long-term sequalae of COVID-19 infection can seriously hinder parent workforce participation. Caregiving as a result of acute illness, long-term consequences, or secondary infections has been a major driver of the gendered socio-economic impacts of the pandemic: Women have experienced higher rates of job loss than their male counterparts, and households headed by single mothers are at significantly increased risk of falling into poverty due to school closures^2-4^.

Implementing multiple overlapping interventions, including masking, improved ventilation, and symptom screening, can reduce viral transmission in school settings^5,6^. Timely detection of infections to enable appropriate isolation of cases, and quarantine or enhanced screening of exposed contacts is crucial for preventing in-school transmission that could lead to larger community outbreaks.

Effective vaccines are now widely available across much of the world. However, in historically marginalized communities, structural barriers that inhibit access to diagnostic testing (i.e., medical mistrust, lack of paid time off, poor geographic access) also present barriers to vaccine uptake^7-9^. Strategies to rapidly identify COVID-19 cases in communities with low testing and vaccine uptake are necessary to achieve health equity, reduce morbidity and mortality, and avoid the emergence of new variants of concern (VoCs) with increased vaccine escape potential^10^.

Moreover, as many have argued from both an ethical and a pragmatic standpoint, there is a need for preferential options for poor communities in which the most promising technological advances are deployed first to where they are needed, rather than to where they can be afforded^11-13^.

Wastewater surveillance has gained attention as a tool for passive surveillance of community- and building-level SARS-CoV-2 infections in municipalities and universities^14^. In California, a large residential university with free, university-mandated testing found that large-scale wastewater monitoring allowed the university to identify cases in specific campus buildings and residential halls. Notifying building occupants following a positive wastewater sample was significantly associated with increased diagnostic testing uptake compared to testing uptake prior to the notification, and 85% of all diagnosed infections among on-campus residents were detected in wastewater^15^.

The passive nature of wastewater sampling is promising for school COVID-19 surveillance in communities where students, parents, and staff are more likely to face structural barriers to vaccination and diagnostic testing uptake. Proof of concept has been previously examined in Kindergarten through 12^th^ grade (K-12) school settings in the UK and the United States^16,17^.

However, two concerns about potential effectiveness of wastewater sampling in these settings are that 1) not all individuals have daily bowel movements on site; and 2) spatial resolution is limited to entire buildings or building clusters because of sewer access locations.

We developed an environmental monitoring system that utilizes wastewater and daily surface sample surveillance to detect COVID-19 cases in elementary schools and childcare settings. We named the project Safer at School Early Alert (SASEA). In this study we report on the accuracy of wastewater and surface sampling within the SASEA program, measured against weekly diagnostic testing, as well as the potential acceptability and efficacy of the program as suggested by staff and student diagnostic testing uptake.

## Methods

We used a prospective observational study design to evaluate the effective accuracy of passive school-based environmental surveillance for detecting the presence of asymptomatic SARS-CoV-2 infection against weekly asymptomatic surveillance diagnostic (PCR) testing. The SASEA project was intended to evaluate the potential real-world utility (effectiveness) of environmental surveillance in socially vulnerable, low resource settings which are arguably entitled to the most scientifically rigorous support to ensure staff and student safety. We worked with stakeholders at the school, district, and county levels to integrate wastewater and surface sampling into school-based public health programming in a way that would be feasible and acceptable for communities with the highest burden of COVID-19 morbidity and mortality.

### Safer at school early alert (SASEA) program and pilot

SASEA consists of four primary components: (1) Daily environmental sampling for SARS-CoV-2 using wastewater from all school restrooms and surface swabs (typically the center of a classroom floor) from individual classrooms; (2) Rapid results reporting to site administrators (approximately 30 hours after sample collection); (3) On-site diagnostic testing of students and staff when SARS-CoV-2 was detected in wastewater or surface samples; and (4) Risk mitigation via environmental modification (e.g., moving classes outdoors, increasing ventilation in classrooms with a potential case) and health communication messaging (e.g., encouraging double masking, recommending wider testing among household members) (Fig. 1).

**Fig 1:**
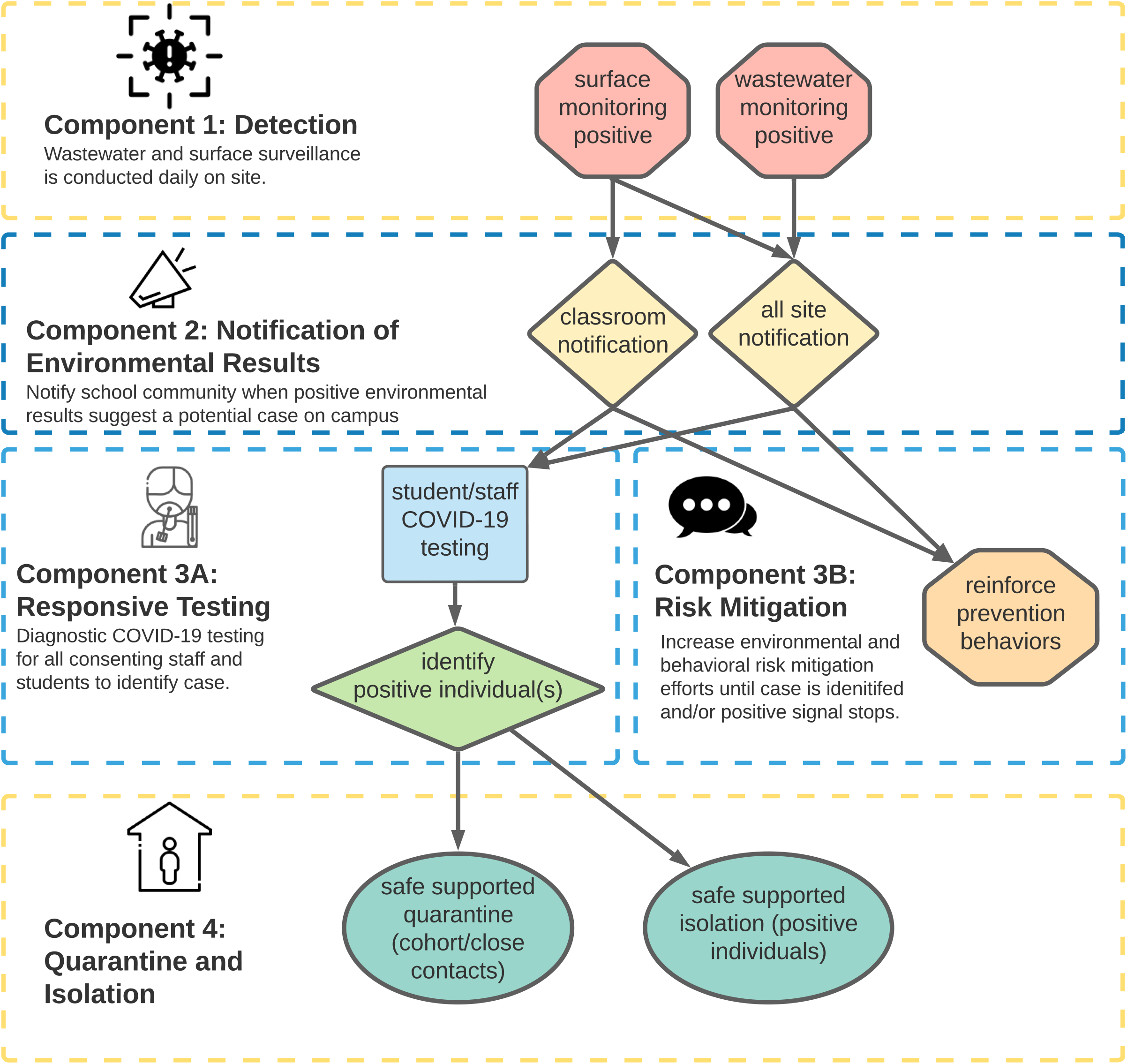
Safer at School Early Alert (SASEA) system.

Surface sampling was included because although SARS-CoV-2 transmission though fomites is uncommon^19^, our team has previously recovered traces of viral RNA by surface sampling of rooms occupied by infected individuals in a hospital setting^20,21^, suggesting that surface sampling can provide a complementary approach to wastewater viral monitoring. The center of the floor was chosen based on previous research conducted by our team which suggests that this is where airborne virus particles are most likely to settle.

We piloted SASEA in nine public elementary schools (grades K-6) in San Diego County, California during the 2020-2021 academic year. Pilot sites were selected from ZIP Codes with COVID-19 rates above the county median and with high levels of social vulnerability according to the California Healthy Places Index (HPI). During a 12-week validation phase (November 16 2020 – March 1, 2021), we conducted daily wastewater monitoring at each site and surface sampling in each classroom where children were present. Approximately 1700 students and staff were regularly present across all nine sites during the validation period. The 7-day average case rate in San Diego County ranged from 6.5 per 100,000 to 69.5 per 100,000 during this phase. Seven-day case rates in our partner schools ranged from 4 cases/100,000 in late February 2021 to a high of 211 per 100,000 in mid-December of 2020.

### Wastewater sample collection

Wastewater autosamplers were installed at each site to collect time-weighted composite samples and programmed to sample every 10-15 minutes over a 7 hour interval (typically 6:30am - 4pm). Sites were sampled each day that children were present (i.e., Monday through Friday excluding school holidays) using Hach autosamplers. Autosamplers were installed at manhole covers or sewage cleanout sites which captured all campus restrooms. The SASEA team worked with school facilities management to ensure that all school wastewater flow was captured at the selected installation point, typically just before the school sewer system joined the main city sewer line. A trained technician collected samples at the end of each school day (2pm - 3:30pm) and transported them to a laboratory at the University of California, San Diego campus. To support the high volume and rapid turnaround necessary for this project, a streamlined, high-throughput wastewater processing pipeline was implemented^22,23^.

The sensitivity of the wastewater processing and detection pipeline was previously validated for building-level resolution as a part of a large study on the UCSD campus where wastewater surveillance was conducted in tandem with clinical surveillance (15, 34). Briefly, SARS-CoV-2 viral RNA was concentrated from 10ml of raw sewage using an automated, high-throughput approach employing affinity capture magnetic hydrogel particles (Ceres Nanosciences Inc., USA). Samples were concentrated in 450uL of lysis buffer and extracted using the MagMax Microbiome Ultra kit (Thermo Fisher Scientific, USA) on the Kingfisher platform. Final elution volume for the extracted nucleic acid was 50uL.

RT-qPCR reactions were run using Promega SARS-CoV-2 RT-qPCR Kit for wastewater (Cat.no. CS317402, Promega, USA). Primer/probe sets targeting the N1, N2, E gene; primers detecting Pepper Mild Mottle Virus (PMMoV) as an internal process control; and an internal amplification control, IAC (for inhibition assessment) were used. A synthetic RNA encoding the E and N genes of SARS-CoV-2 was used as the positive control. A ladder of 6-fold dilutions was run with every RT-qPCR run (Promega Cat. #CS317402). The no-template control (NTC) was nuclease-free water. Additionally, to detect inhibition in samples, the positive control RNA ladder was run with every RT-qPCR run (5 1:10 serial dilutions of a positive control). We did not find any significant differences for dilutions in nuclease free-water (verified to be SARS-CoV-2 negative) spiked in with the same dilutions. Cq values for the no dilution to 1:100000 dilution were not significantly different, suggesting no PCR inhibition in the RNA extracts. Two-tailed t tests at 95% confidence interval were used to determine if the average Cq values were statistically significant from the spiked-in water control. SARS-CoV-2 RNA were quantified as genome equivalents per liter (GE/L) by deriving the linear regression between the log10(GE) and Cq value of a standard curve comprising an eightfold serial dilution of heat-inactivated SARS-CoV-2 viral particles and normalizing for RT-qPCR input material and sample volume. Four technical replicates were performed per dilution. Limits of detection (LODs) at 95% confidence were defined as gene equivalents where amplification in all replicates was observed^22^.

### Surface sample collection

Surface samples were collected daily from all classrooms with stable cohorts (i.e., classrooms used for brief one-on-one services for students were excluded). Classroom teachers and/or custodial staff swabbed a one-foot square area in the center of each classroom floor at the end of each day prior to classroom cleaning. Additional details about surface sampling methodology and technical performance are available elsewhere^24^. The principal and COVID-19 liaison were notified of wastewater or surface sample results by email (typical turnaround timebetween 26 and 36 hours). All sites were given template language to notify staff and parents of positive results, although sites chose to implement these notifications in a variety of ways. Sites were also given educational materials on contact tracing, diagnostic testing access, and supportive services available for individuals who tested positive.

### Cost Estimation

We estimated the one-time start-up costs and weekly recurrent costs of the wastewater and surface sample collection per school from a payer perspective using a micro-costing approach. Start-up costs included: training, personnel time for installation of the wastewater sampler and development of training materials, the wastewater sampler, and additional equipment associated with wastewater sampler installation. Recurrent costs included: personnel costs for sample collection and transport, mileage, and lab processing fees. Results are provided in 2020 USD. An additional analysis estimated the costs of a wastewater only or surface sampling only program.

### Reference standard

We validated the wastewater and surface samples against weekly diagnostic testing for all consenting students and staff on campus. All individuals who enrolled in weekly diagnostic testing provided written informed consent (staff) or parental consent and participant assent (students). Typical turnaround time from sample collection to results notification was 24 hours or less, and so for individuals who tested positive, the date of testing was also typically the last day on campus.

Diagnostic anterior nasal swab samples were collected once a week from all consented students and staff by trained healthcare personnel (certified nursing assistants, phlebotomists, or emergency medical technicians). Samples were processed at the University of California, San Diego EXCITE CLIA laboratory and tested for the presence of the SARS-CoV-2 virus with an RT-qPCR assay based on a miniaturized version of the ThermoFisher COVID-19 detection kit (PN: A47814, ThermoFisher, Carlsbad, CA) under an FDA EUA.

The County of San Diego, Health and Human Services Agency (SDHHSA) agency shared de-identified data for all cases that had been present at any of our pilot sites. We worked with school principals and COVID-19 liaisons to match each case to a classroom. School administrators provided additional information about cases that were reported to them, including the last known date the individual was on campus.

### Statistical validation of environmental samples against reference standard

We assessed concordance between environmental samples and diagnostic testing by determining how often a positive diagnostic test preceded or coincided with a positive environmental sample (retrospective analysis) and conversely, how often a positive environmental sample coincided or was followed by a positive diagnostic test (prospective analysis). Under the retrospective analysis, the unit of analysis was positive diagnostic tests, and the retrospective outcome was the proportion of diagnostics tests preceded/coincided, within 7-days, by a positive wastewater or positive surface sample. A 7-day window was utilized for these analyses based on the frequency of onsite surveillance testing being offered by the study team. Under the prospective analysis, the unit of analysis was positive wastewater samples and positive surface samples, and the prospective outcome was the proportion of environmental samples coincided/followed, within 7-days, by a positive diagnostic test. We computed 95% confidence intervals for the retrospective and prospective outcomes by randomly sampling 10,000 times from independent normal distributions for the two proportions. Analysis was limited to sites that participated in the SASEA program throughout the validation phase and whose diagnostic test could be linked back to a classroom (for assessing concordance between diagnostic test and surface samples). All analyses were conducted in STATA version 16.

### Viral sequencing

Next generation sequencing libraries from the SARS-CoV-2 positive diagnostic (anterior nares) samples were prepared using a miniaturized version of the Swift Normalase® Amplicon Panel kit (PN: SN-5×296 (core) COVG1V2-96 (amplicon primers), Integrated DNA Technologies, Coralville, IA) and sequenced on the NovaSeq 6000 platform.

Surface and wastewater data were analyzed with the COVID-19 VIral Epidemiology Workflow (C-VIEW). C-VIEW, available at https://github.com/ucsd-ccbb/C-VIEW, is an open-source, end-to-end workflow for viral epidemiology that is currently focused on SARS-CoV-2 lineage assignment and phylogenetics. Starting from raw sequencing data (.fastq) files, it performs alignment, variant identification, consensus sequence calling, lineage assignment, phylogenetic tree building, and calculation of extensive quality control metrics.

Clinical and environmental samples that were positive for SARS-CoV-2 viral RNA via qPCR were sequenced using the Swift Normalase® Amplicon Panels (SNAP) kit. A miniaturized version of the protocol was implemented for library generation and indexed using dual indexing oligos (SN91384) which yielding up to 1536 index pairs per pool. Details on the library generation protocol are provided elsewhere by Karthikeyan et al^25^.

Positive and negative controls were included for all stages of sample processing including sequencing. In case any of the controls failed or indicated cross-contamination, the entire batch was rerun. Clinical and wastewater samples were processed separately during sequencing due to significant differences in viral load between the two sample types. 95.3% of the sequences passed QC threshold of at least 70% SARS-CoV-2 genome coverage with no evidence of cross-contamination as well as positive and negative controls passing QC for the run Libraries were pooled with equal volume and sequenced with Single Read 26 basepairs (SR26) on an Illumina MiSeq using a MiSeq Reagent Nano Kit V2 to determine volumes for balanced loading.

### Ethical considerations

All study procedures were reviewed and approved by the University of California, San Diego Institutional Review Board (IRB). The IRB determined that wastewater and surface samples were not human subjects data. Weekly diagnostic testing of students and staff was determined to be minimal risk, (i.e., not more than the typical risk of similar weekly diagnostic testing being offered in a variety of school settings at the time), and the whole protocol underwent full IRB review (protocol 201664). Linkage of the environmental samples to the human sample information was also covered by protocol 201664. All participants provided informed consent (if 18 or above) or assent plus parental consent (if under the age of 18) to engage in diagnostic testing. The study team only retained de-identified diagnostic outcome data (sample result, classroom) for the purposes of analyses. The types of ethical considerations we included for the human specimens (clinical nasal swabs, not fecal specimens), included possible discomfort to the subject, possible incidental findings about the subject’s microbiome or infection status if the specimens were used for assays beyond SARS-CoV-2, embarrassment during the swabbing procedure or when being asked specific questions on the questionnaire, and the need for secure storage of the forms and data to prevent accidental data disclosure.

## Results

Consent to participate in weekly diagnostic testing rose steadily throughout the 12-week study period. By week 12, 1,294 (75.4%) of the 1,717 individuals consistently present at the sites had consented to onsite diagnostic testing, of which 1,275 (98.5%) were tested at least once.

Approximately 25% of people on campus on a given day were staff and 75% were students. Staff had an overall consent rate of 99.3% by the end of the study period, compared to 60% consent for students. Median student age was 8.5 years (range: 0 – 17.5 years). Median staff age was 42.5 (range: 18 – 78). Among students, 37.6% were female, 39.8% were male, and 22.6% of parents declined to provide information related to gender. Among staff, 67.3% of consented individuals were female, 14.2% were male, and 18.6% declined to state. Approximately 23% of students and 34% of staff identified as white and non-Hispanic, 63% of students and 54% of staff identified as Hispanic, and 6% of students and 3% of staff identified as Black or African American.

There were 447 data collection days across the nine sites (i.e., approximately 50 school days per site over the 12-week study period). In this period, SARS-CoV-2 was detected in 374 surface samples and 133 wastewater samples. Eighty-nine individuals tested positive; 42 via onsite testing and 47 through outside testing. We do not have data on the number of outside tests among students or staff that received negative results.

Of the 89 identified on-campus cases, 83 (93%) were preceded by a positive wastewater or same-room surface sample in the 7-day window preceding the individual’s last day on campus (95% CI: 88% - 98%). The majority of these, 60 (67%), were associated with a positive wastewater sample (95% CI: 57% - 77%). Of the 72 identified cases among individuals associated with a single classroom, 29 (40%) corresponded with a positive surface sample in the associated room in the 7-day window preceding the individual’s last day on campus (95% CI: 29% - 52%).

Positive surface or wastewater signals occurred on 240 (60%) of study days, during which 76 (28%) days had a positive wastewater and surface signal on the same day. Just under half (47%, n=127) of days with positive signals were associated with a diagnosed case in the 7-day window following the signal (95% CI: 41% - 52%). Seventy (53%) of positive wastewater signals were followed by an identified case within 7 days (95% CI: 44% - 61%), while 40 (11%) of positive surface samples were followed by a detected case within 7 days (95% CI: 8% - 14%).

By week 9, we had obtained consent for 70% of eligible students and staff across all sites. In weeks 9-12, there were 157 positive surface samples, 67 positive wastewater samples, and 19 identified cases, 15 of which were associated with a classroom (the remaining 4 cases were among non-classroom staff members). Nine of the cases (60%) linked to a classroom were associated with a positive surface sample (95% CI: 36% - 85%) and 18 identified cases (95%) were associated with a positive wastewater sample (95% CI: 85% - 100%). In the same time period, positive wastewater or surface samples occurred on 130 days and 44 (34%) of these were associated with an identified case (95% CI: 26% - 42%). All 19 cases (100%) identified in this were associated with either a surface sample, wastewater sample, or both (figure 2).

**Fig 2:**
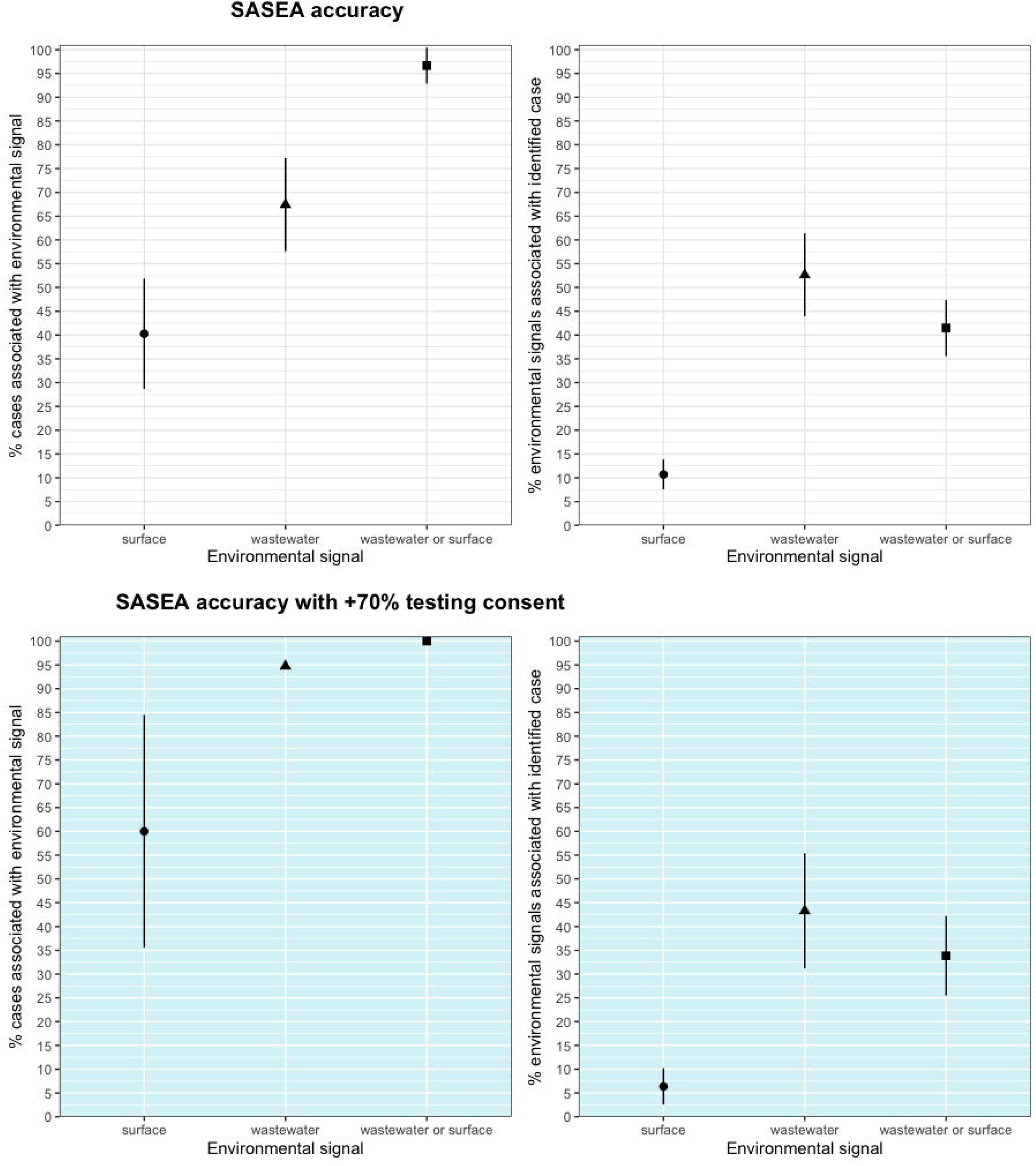
Wastewater and surface sampling and 95% confidence interval across full 12-week pilot period, and with consent at 70% or above (weeks 9-12)

Testing uptake within SASEA partner schools was higher than in nearby districts. In February of 2021, approximately 13% of onsite students and staff in a large local district accessed onsite COVID-19 tests^26^, compared to 78% of students and staff across all SASEA partner sites in the same time period. Five schools achieved consent rates over 90%, and at 3 sites 100% of all on-campus staff and students had consented to weekly testing by the end of the validation period (figure 3).

**Fig 3:**
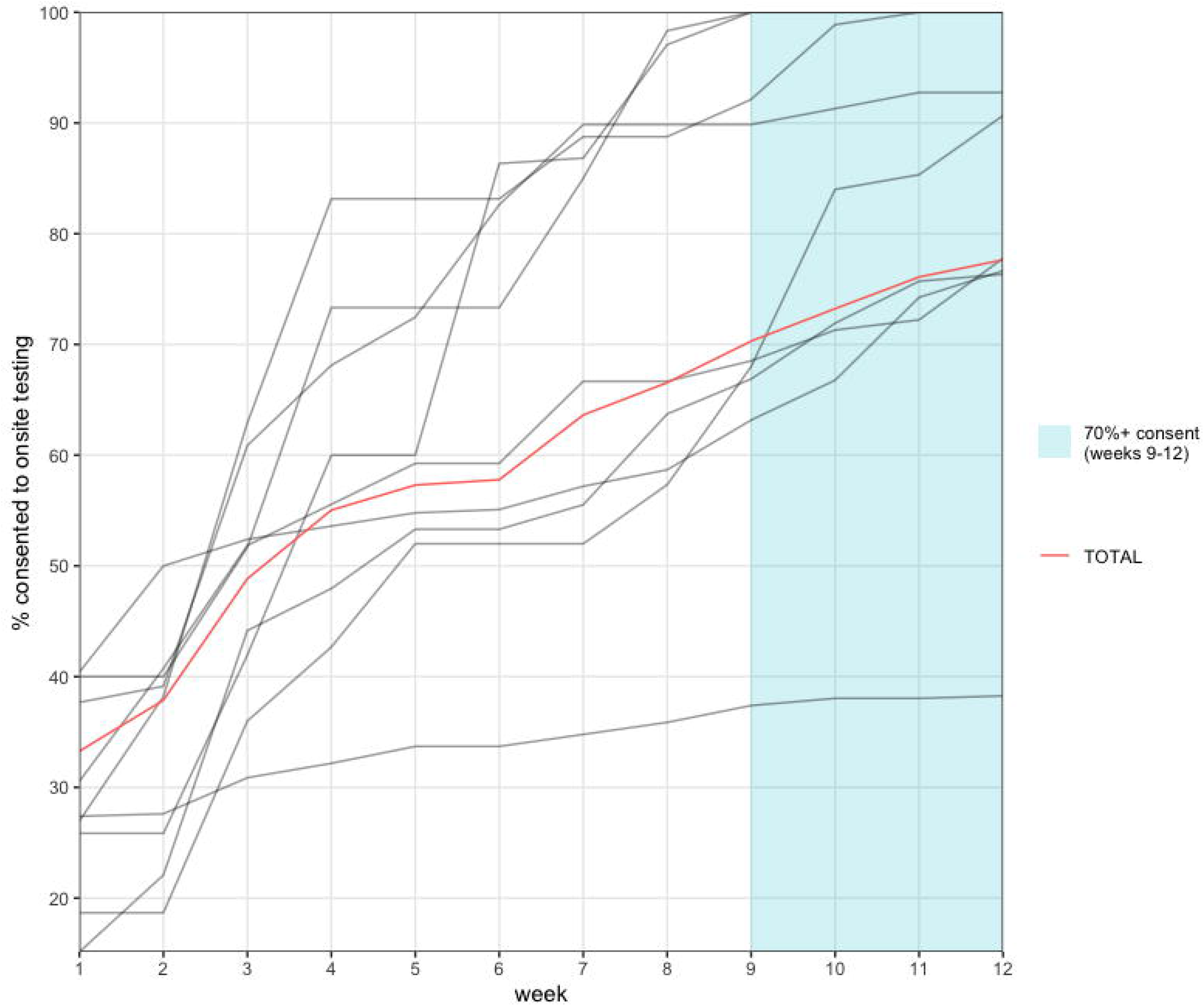
Individual diagnostic testing consent over time by site (gray) and total across all 9 sites (red) throughout 12-week pilot phase.

### Cost estimation for wastewater and surface sampling collection

The one-time start-up cost was $4,228 per school ($3,789 for wastewater start-up, $439 for the surface sample start-up). The cost of the wastewater sampler ($3,200) comprised the bulk of the start-up costs. The recurrent costs were $2,745 per week per school, of which the majority (71%) was the sample processing cost. We estimated weekly recurrent costs of $892/week for a wastewater sampling only program, and $2,136 for surface sampling only program, with small efficiencies observed by running both programs together. Assuming 180 school days (36 weeks) and 300 students, the total cost works out to just under $350 per student per year. Limiting the system to wastewater surveillance alone would cost approximately $120 per student per year in a school with 300 students.

### Sequencing

Sixty-four of 133 positive wastewater samples yielded near-complete SARS-CoV-2 viral genomes (average genome coverage of 93.2%). Ninety-five percent of the sequenced samples passed QC-at least 70% of SARS-CoV-2 genome coverage with no evidence of cross-contamination as well as positive and negative controls passing QC for the run^26^. Sequencing of the environmental samples enabled recovery of near complete SARS-CoV-2 genomes (>99% genome coverage) from wastewater samples with cycle threshold values as high as 37.6 using a miniaturized tiled amplicon sequencing approach^25^. All positive surface samples submitted for tiled amplicon sequencing (n=10) generated near-complete viral genomes (genome coverage > 98%).

One SARS-CoV-2 genome sequenced from a carpeted floor surface was associated with a genome from a SASEA clinical testing sample via clustering in a phylogenetic tree (figure 4A). The individual whose diagnostic sample was sequenced was confirmed to have been present in the classroom with a positive surface sample.

**Figure 4:**
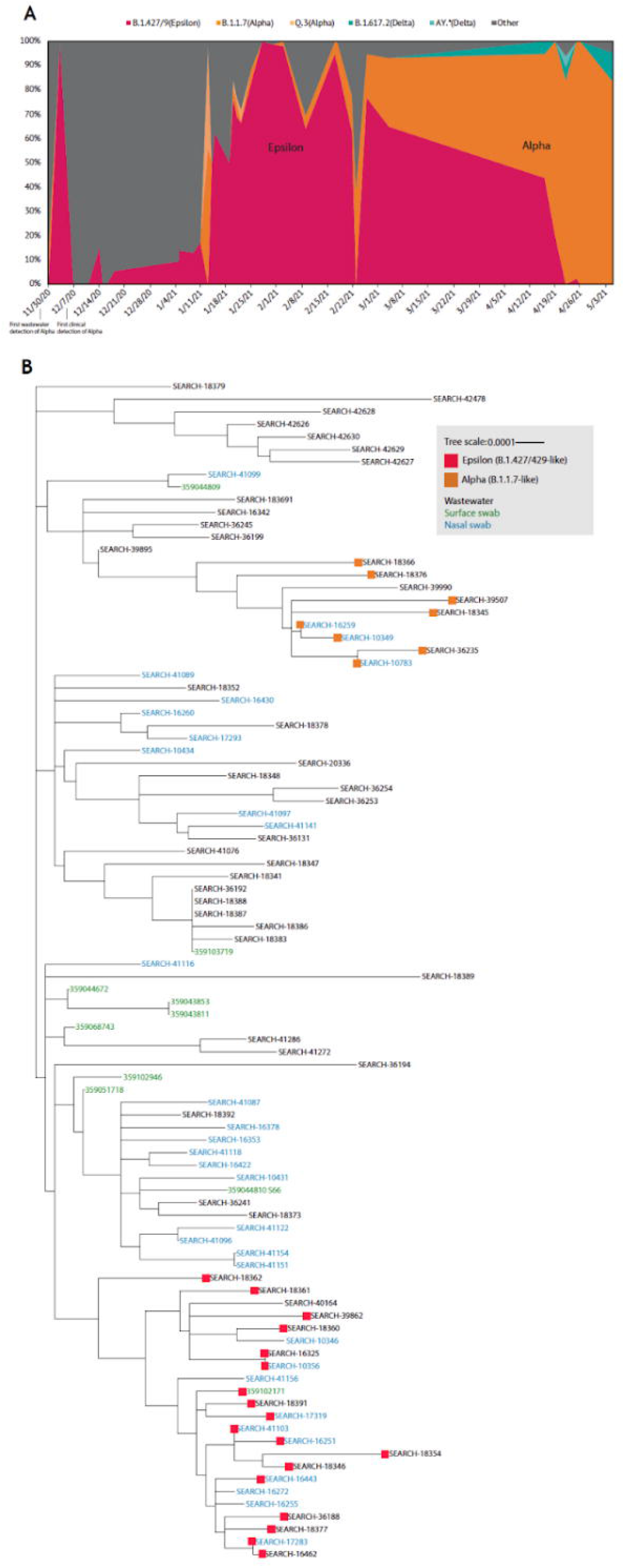
SARS-CoV-2 genomic surveillance. A. SARS-CoV-2 variant prevalence in the wastewater sequences: The relative proportions of each variant were calculated using Freyja v.1.3.11^25^ ‘Other’ contains all lineages not designated as VOC/VOIs B. Genomic sequencing of clinical and environmental samples: Maximum likelihood (ML) phylogenetic tree for the clinical and environmental (wastewater and surface) samples which had an average SARS-CoV-2 virus genome coverage of 95% or above constructed using IQtree.

Among the sequenced samples, we identified the Alpha variant (B.1.1.7) in 14.2% of the wastewater samples and 8.6% of SASEA diagnostic tests (nasal swabs) and the Epsilon variant (B.1.427/B.1.429) in 22.2% of wastewater samples, 25.0% of SASEA clinical tests, and 10.0% of surface samples (figure 4B). When sequencing data was available for both environmental monitoring modalities with matched spatial-temporal characteristics (same school, 5-day time window), we were able to match strain identifications between surface and wastewater positive samples. The Alpha variant was detected in a November 30th wastewater sample, 30 days before it was first identified in the region. Following the pilot phase, the Delta variant (B.1.617.2) was identified at a school site on April 16th, 5 days after it was first identified in the county via diagnostic testing.

## Discussion

Our findings suggest that environmental surveillance via wastewater and surface sampling can be an effective passive screening tool to complement and potentially enhance individual testing approaches. We found high concordance between diagnosed cases and positive environmental samples, with lower concordance between positive environmental samples and diagnosed cases. Diagnostic testing consent is crucial for system effectiveness; however, it is important to acknowledge that parents and staff may not consent to onsite diagnostic testing for a wide array of reasons. Positive environmental signals should prompt the increased use of risk mitigation measures (i.e., masking/double-masking, social distancing, ventilation) while waiting for responsive testing implementation and results, and/or in the absence of identifying a case.

Our findings should be interpreted with its limitations in mind. Our observation that SASEA sites had higher participation in weekly diagnostic testing than comparable nearby schools was consistent with other settings where environmental monitoring notifications have been shown to increase diagnostic testing uptake^15^. However, more research is needed to understand how other aspects of our study design, including the requirement to consent to testing as part of an ongoing research process, recruitment strategies and school administrator involvement, influenced school-wide testing rates. SDHHSA provided data on cases affiliated with SASEA partner sites, but our denominator only includes known campus-associated cases.

We lack data on cases that may have contributed to surface or wastewater samples during our 12-week validation phase but did not test through SASEA or an agency that reports to SDHHSA. San Diego has close social and economic links with Tijuana, Mexico and four of our partner schools were within 10 miles of the San Ysidro Port of Entry. SDHHSA authorities are notified of all cases in other counties or countries (i.e., Mexico) that provide a residential address in San Diego County. However, it is possible that some individuals may have tested elsewhere without providing a residential address in the county. It is also possible that individuals may have tested positive using an at-home test kit and declined to notify SDHHSA of positive results. We also do not have data on individuals who tested and received negative results through outside providers or at home test kits during our 12-week pilot phase. For these reasons we are unable to calculate the true sensitivity and specificity of the environmental monitoring system. However, to our knowledge, ours is the only systematic investigation of the accuracy of environmental monitoring to detect and rapidly respond to cases in school sites.

Additionally, although we present evidence for the utility of the SASEA system and the cost of the intervention, we note that the costs presented are based on the availability of a lab which could perform high-volume, rapid turnaround processing of SARS-CoV-2 wastewater and surface samples at the scale implemented. Further scale-up could require additional laboratory equipment and personnel not considered in this analysis. Additionally, sample processing costs will likely vary across laboratories, and some schools may reside in areas of the country without access to laboratories able to provide the rapid turnaround as in our study, raising important questions of equity.

The US Centers for Disease Control (CDC) has supported the rapid scale-up of wastewater surveillance for SARS-CoV-2 through the National Wastewater Surveillance System (NWSS). The NWSS focuses on relatively large regions served by wastewater treatment plants^27^. School-based environmental surveillance can augment these large regional efforts in three ways: First, through genomic surveillance with a higher level of spatial resolution, allowing for more rapid detection and targeted response to emergent VoCs. Second, by providing neighborhood level data, allowing public health officials to deploy finite resources more precisely. And third, to encourage school community members to access individual level testing when environmental surveillance data suggests it is most necessary.

As rapid antigen testing (RATs) becomes more commonplace among individuals, environmental monitoring has become an increasingly important tool for genomic surveillance and rapid identification of new variants of concern (VOC). Moreover, while wastewater detection of SARS-CoV-2 viral RNA via PCR-based approaches is valuable in tracking the viral prevalence at a municipal level, viral genome sequencing of positive wastewater samples can help elucidate strain geospatial distributions, thereby aiding in identification of outbreak clusters and more targeted tracking of prevailing or newly emerging variants. The methods used in this study to detect SARS-CoV-2 viral RNA allowed us to sequence viral genomes directly from the wastewater to characterize the circulating viral variants/lineages. This technique may have applications in areas where sequencing capacity and/or individual diagnostic testing uptake is limited. Similarly, SARS-CoV-2 genomes sequenced from wastewater can be associated to nasal samples via clustering, while consecutive observations of specific genomes in wastewater suggest persistent viral shedding.

In the United States, neighborhood public schools serve specific geographic regions. As a result, environmental monitoring has the additional benefit of acting as an early warning system for the local community from which students are drawn. Utilizing public schools as community sentinel surveillance sites for SARS-CoV-2 can support community-tailored interventions such as ensuring materials are translated into languages spoken in the community, working with field staff to ensure culturally competent outreach, and providing community-specific testing and vaccine clinics. School-based wastewater epidemiological surveillance provides more spatial granularity than municipal wastewater treatment, while allowing school communities to provide specific, timely, and tailored risk mitigation advice to parents, students, and staff.

While we pilot-tested SASEA in relatively small elementary schools during a time of limited in-person attendance, the system has even greater potential to be cost-effective in larger school settings such as middle and high schools. Because students frequently change classrooms in middle and high schools surface sampling would be of limited utility. The average high school in the United States has approximately 850 students. At just under $36,000 per year, a year of passive wastewater surveillance in this setting would be less than $50 per student per year, equivalent to less than half of one diagnostic PCR test^28^, with nearly equivalent accuracy and entire school coverage.

The SASEA system was designed for communities that face social and structural barriers to diagnostic testing and vaccine access. These barriers are likely to become more pronounced with the impending end of the federal COVID-19 emergency declaration in the United States, which will severely reduce access to RATs and diagnostic testing^29^. Wastewater and surface sample monitoring are anonymous, aggregate, and can provide sentinel surveillance for early detection of outbreaks and VoCs. Even in the absence of a diagnosed case, positive environmental samples serve as a behavioral cue to increase or re-implement risk mitigation measures in a classroom or entire school.

## Data Availability

School level environmental, diagnostic, and consent data have been archived in the Dataverse repository and are available at https://doi.org/10.7910/DVN/HD62J4. All wastewater and nasal-swab sequencing data have been deposited to GISAID and their accession details provided in Supplemental File Data S1

## Acknowledgments

We are deeply grateful to the leadership, staff, students, and families at our partner school districts and school sites across San Diego County. This project would not have been possible without their enthusiasm and generosity. We thank our colleagues at the County of San Diego, Health and Human Services Agency for their vision and support, particularly John Malone, Sarah Stous, and Rorick Luepton. At UCSD, Isabella Cuturrofo, Jeanessa Mendoza, Kristine Ngo, Jessica Ni, and Elizabeth Frost all provided vital support with project implementation. Austin Dahl provided extensive support with data management and visualization. RFM thanks Ms. Esther J. Krohne for inspiring the project.

The opinions and assertions expressed herein are those of the authors and do not necessarily reflect the official policy or position of the County of San Diego Health and Human Services Agency

## Author Contributions

RFM: Conceptualization; Methodology; Formal Analysis; Investigation; Writing - original draft; Visualization; Supervision; Funding Acquisition

SK: Methodology; Formal Analysis; Investigation; Data curation; Writing - original draft; Visualization

TG: Methodology; Formal Analysis; Writing - original draft.

RSG: Methodology; Data curation; Writing - original draft; Visualization; Supervision.

RS: Methodology; Formal Analysis; Investigation; Writing - original draft.

VC: Methodology; Formal Analysis; Investigation; Data curation; Writing - original draft.

LK: Conceptualization; Methodology; Investigation; Data curation; Writing - original draft; Supervision; Project administration; Funding Acquisition

NKM: Methodology; Writing - original draft.

AW: Methodology; Data curation; Formal Analysis; Writing – revised draft

CW: Supervision; Project administration.

MF: Investigation; Data curation.

VO: Investigation; Data curation.

AM: Investigation; Data curation.

PGZ: Investigation; Supervision.

MN: Investigation; Data curation.

AVV: Investigation; Data curation; Visualization

TL: Investigation.

DD: Investigation.

AH: Investigation.

ST: Resources; Supervision; Project administration.

KJ: Methodology; Investigation; Resources.

BH: Methodology; Investigation.

AH: Methodology; Investigation.

AB: Methodology; Investigation.

AMM: Methodology; Investigation.

CAN: Methodology; Investigation.

SBR: Methodology; Investigation.

NM: Methodology; Investigation.

KMF: Methodology; Investigation.

GH: Investigation; Data curation.

SF: Investigation; Data curation.

HMT: Investigation; Data curation.

TV: Investigation; Data curation.

JM: Investigation; Data curation.

JK: Investigation; Data curation.

BK: Investigation; Data curation.

CY: Investigation; Data curation.

ADA: Investigation; Data curation.

SE: Investigation; Data curation.

JE: Investigation; Data curation.

KC: Investigation; Data curation.

LCL: Resources

TR: Investigation; Data curation.

RK: Conceptualization; Methodology; Resources; Writing - original draft; Supervision.

## List of supplementary materials

Data S1: Wastewater and nasal-swab sequencing data GISAID accession details

S2: Costing methods and expanded results

## REFERENCES

1. N. Kalluri, C. Kelly, A. Garg, Child care during the COVID-19 pandemic: A bad situation made worse. Pediatrics >147, (2021).

2. R. J. Petts, D. L. Carlson, J. R. Pepin, A gendered pandemic: Childcare, homeschooling, and parents’ employment during COVID□19. Gender, Work & Organization 28, (2020).

3. M. I. Cardel, N. Dean, D. Montoya-Williams, Preventing a secondary epidemic of lost early career scientists. Effects of COVID-19 pandemic on women with children. Annals of the American Thoracic Society 17, 1366–1370 (2020).

4. T. Alon, M. Doepke, J. Olmstead-Rumsey, M. Tertilt, “The impact of COVID-19 on gender equality,” (National Bureau of economic research, 2020).

5. J. Lessler, M. K. Grabowski, K. H. Grantz, E. Badillo-Goicoechea, C. J. E. Metcalf, C. Lupton-Smith, A. S. Azman, E. A. Stuart, Household COVID-19 risk and in-person schooling. Science 372, 1092–1097 (2021).

6. J. Gettings, M. Czarnik, E. Morris, E. Haller, A. M. Thompson-Paul, C. Rasberry, T. M. Lanzieri, J. Smith-Grant, T. M. Aholou, E. Thomas, C. Drenzek, D. MacKellar, Mask Use and Ventilation Improvements to Reduce COVID-19 Incidence in Elementary Schools.MMWR Morb Mortal Wkly Rep 70, 779–784 (2021).

7. L. Doornekamp, L. van Leeuwen, E. van Gorp, H. Voeten, M. Goeijenbier, Determinants of Vaccination Uptake in Risk Populations: A Comprehensive Literature Review. Vaccines 8, p(2020).

8. C. Dodds, I. Fakoya, Covid-19: ensuring equality of access to testing for ethnic minorities. BMJ 369, m2122 (2020).

9. N. E. MacDonald, R. Butler, E. Dube, Addressing barriers to vaccine acceptance: an overview. Human Vaccines & Immunotherapeutics 14, 218–224 (2018).

10. A. Fontanet, B. Autran, B. Lina, M. P. Kieny, S. S. Abdool Karim,SARS-CoV-2 variants and ending the COVID-19 pandemic. The Lancet 397, 952–954 (2021).

11. J. P. Figueroa, P. J. Hotez, C. Batista, Y. B. Amor, O. Ergonul, S. Gilbert, M. Gursel, M. Hassanain, G. Kang, D. C. Kaslow, J. H. Kim, B. Lall, H. Larson, D. Naniche, T. Sheahan, S. Shoham, A. Wilder-Smith, S. O. Sow, N. Strub-Wourgaft, P. Yadav, M. E. Bottazzi, Global public health security and justice for vaccines and therapeutics in the COVID-19 pandemic. EClinicalMedicine 39, 101053 (2021).

12. J. C. G. Corpuz, The vulnerable in time of pandemic: toward a preferential option for the vulnerable and marginalized. Journal of Public Health fdab253 (2021).

13. A. Martins, Ethics and Equity in Global Health: The Preferential Option for the Poor. Journal of Moral Theology 1(CTEWC Book Series 1), 96–105 (2021).

14. F. Hassard, L. Lundy, A. C. Singer, J. Grimsley, M. Di Cesare, Innovation in wastewater near-source tracking for rapid identification of COVID-19 in schools. The Lancet Microbe 2, e4–e5 (2021).

15. S. Karthikeyan, A. Nguyen, D. McDonald, Y. Zong, N. Ronquillo, J. Ren, J. Zou, S. Farmer, G. Humphrey, D. Henderson, T. Javidi, K. Messer, C. Anderson, R. Schooley, N.K. Martin, R. Knight, Rapid, Large-Scale Wastewater Surveillance and Automated Reporting System Enable Early Detection of Nearly 85% of COVID-19 Cases on a University Campus. mSystems 6, e00793–21 (2021).

16. J. Crowe, A. T. Schnaubelt, S. SchmidtBonne, K. Angell, J. Bai, T. Eske, M. Nicklin, C. Pratt, B. White, B. Crotts-Hannibal, N. Staffend, V. Herrera, J. Cobb, J. Conner, J. Carstens, J. Tempero, L. Bouda, M. Ray, J. V. Lawler, W. S. Campbell, J.-M. Lowe, J. Santarpia, S. Bartelt-Hunt, M. Wiley, D. Brett-Major, C. Logan, M. J. Broadhurst, Assessment of a Program for SARS-CoV-2 Screening and Environmental Monitoring in an Urban Public School District. JAMA Netw Open 4, e2126447 (2021).

17. V. C. Gutierrez, F. Hassard, M. Vu, R. Leitao, B. Burczynska, D. Wildeboer, I. Stanton, S. Rahimzadeh, G. Baio, H. Garelick, J. Hofman, B. Kasprzyk-Hordern, R. Kwiatkowska, A. Majeed, S. Priest, J. Grimsley, L. Lundy, A. C. Singer, M. D. Cesare, Monitoring occurrence of SARS-CoV-2 in school populations: a wastewater-based approach. https://doi.org/10.1101/2021.03.25.21254231 (2021).

18. A. C. Feldstein, R. E. Glasgow, A Practical, Robust Implementation and Sustainability Model (PRISM) for Integrating Research Findings into Practice. The Joint Commission Journal on Quality and Patient Safety 34, 228–243 (2008).

19. E. A. Meyerowitz, A. Richterman, R. T. Gandhi, P. E. Sax, Transmission of SARS-CoV-2: A Review of Viral, Host, and Environmental Factors. Ann Intern Med 174, 69–79 (2021).

20. C. Marotz, P. Belda-Ferre, F. Ali, P. Das, S. Huang, K. Cantrell, L. Jiang, C. Martino, R. E. Diner, G. Rahman, D. McDonald, G. Armstrong, S. Kodera, S. Donato, G. Ecklu-Mensah, N. Gottel, M. C. Salas Garcia, L. Y. Chiang, R. A. Salido, J. P. Shaffer, M. Bryant, K. Sanders, G. Humphrey, G. Ackerman, N. Haiminen, K. L. Beck, H. -C. Kim, A. P. Carrieri, L. Parida, Y. Vázquez-Baeza, F. G. Torriani, R. Knight, J. Gilbert, D. A. Sweeny, S. M. Allard, SARS-CoV-2 detection status associates with bacterial community composition in patients and the hospital environment. Microbiome 9, 132 (2021).

21. V.J. Cantú, R. A. Salido, S. Huang, G. Rahman, R. Tsai, H. Valentine, C. Magallanes, S. Aigner, N. Baer, T. Barber, P. Belda-Ferre, M. Betty, M. Bryant, M. Casas Maya, A. Castro-Martínez, M .Chacón, W. Cheung, E. Crescini, P. De Hoff, E. Eisner, S. Farmer, A. Hakim, L. Kohn, A. Lastrella, E. Lawrence, S. Morgan, T. Ngo, A. Nouri, R. Ostrander, A. Plascencia, C. Ruiz, S. Sathe, P. Seaver, T. Schwartz, E. Smoot, T. Valles, G. Yeo, L. Laurent, R. Fielding-Miller, R. Knight. SARS-CoV-2 Distribution in Residential Housing Suggests Contact Deposition and Correlates with Rothia sp. mSystems. (2022).

22. S. Karthikeyan, N. Ronquillo, R. Belda-Ferre, D. Alvarado, T. Javidi, C. A. Longhurst, R. Knight, High-Throughput Wastewater SARS-CoV-2 Detection Enables Forecasting of Community Infection Dynamics in San Diego County. mSystems 6, e00045–00021 (2021).

23. C. H. Sheikhzadeh, L. Gonzalez-Barranca, A.M. Adams, O. Ott, S. Karthikeyan, L. Marotz, G. Humphrey, dx.doi.org/10.17504/protocols.io.bshvnb66 (2021).

24. R. A. Salido, V. J. Cantú, A. E. Clark, S. L. Leibel, A. Foroughishafiei, A. Saha, A. Hakim, A. Nouri, A. L. Lastrella, A. Castro-Martínez, A. Plascencia, B. Kapadia, B. Xia, C. Ruiz, C. A. Marotz, D. Maunder, E. S. Lawrence, E. W. Smoot, E. Eisner, E. S. Crescini, L. Kohn, L. F. Vargas, M. Chacón, M. Betty, M. Machnicki, M. Y. Wu, N. A. Baer, P. Belda-Ferre, P. De Hoff,, P. Seaver, R. T. Ostrander, R. Tsai, S. Sathe, S. Aigner, S. C. Morgan, T. T. Ngo, T. Barber, W. Cheung, A. F. Carlin, G. W. Yeo, L. C. Laurent, R. Fielding-Miller, R. Knight, Comparison of heat-inactivated and infectious SARS-CoV-2 across indoor surface materials shows comparable RT-qPCR viral signal intensity and persistence. mSystems. https://doi.org/10.1101/2021.07.16.452756 (2021).

25. Karthikeyan S, Levy JI, De Hoff P, Humphrey G, Birmingham A, Jepsen K, Farmer S, Tubb HM, Valles T, Tribelhorn CE, Tsai R. Wastewater sequencing reveals early cryptic SARS-CoV-2 variant transmission. Nature. 2022 Sep;609(7925):101–8.

26. “San Diego Unified School District COVID-19 School Testing Results.” San Diego Unified School District, San Diego, CA, 25 August 2021, https://web.archive.org/web/20210812134829/ https://sandiegounified.org/covid-19_status/covidtesting

27. Kirby AE, Walters MS, Jennings WC, Fugitt R, LaCross N, Mattioli M, Marsh ZA, Roberts VA, Mercante JW, Yoder J, Hill VR. Using wastewater surveillance data to support the COVID-19 response—United States, 2020–2021. Morbidity and Mortality Weekly Report. 2021 Sep 9;70(36):1242.

28. Kurani N, Pollitz K, Cotliar D, Ramirez G, Cox C. COVID-19 test prices and payment policy [Internet]. Kaiser Family Foundation; 2021 Apr [cited 2022 Dec 12]. (Health Spending). Available from: https://www.healthsystemtracker.org/brief/covid-19-test-prices-and-payment-policy/

29. Cubanski J, Kates J, Guth M, Pollitz K, Musumeci M, Freed M. What Happens When COVID-19 Emergency Declarations End? Implications for Coverage, Costs, and Access [Internet]. Kaiser Family Foundation; 2022 Apr [cited 2022 Dec 12]. Available from: https://www.kff.org/coronavirus-covid-19/issue-brief/what-happens-when-covid-19-emergency-declarations-end-implications-for-coverage-costs-and-access/

